# A Common *CD36* Variant and the Genetic Landscape of Dilated Cardiomyopathy in Individuals of African Ancestry

**DOI:** 10.64898/2026.05.26.26353838

**Authors:** Saketh Challa, Kiran Biddinger, Sarah Abramowitz, Alice Zheng, Jonathan O. Mead, Renae L. Judy, Sean Jurgens, Liam Gaziano, Xin Wang, Seung Hoan Choi, Jennifer Halford, Elizabeth Jordan, Joseph Liu, VA Million Veteran Program, Penn Medicine Biobank, Kyong-Mi Chang, Amanda Vest, W.H. Wilson Tang, Philip Tsao, Daniel D. Kinnamon, Scott M. Damrauer, Patrick T. Ellinor, Michael Levin, Ray E. Hershberger, Jennifer E. Huffman, Krishna G. Aragam

## Abstract

**Importance:** Dilated cardiomyopathy (DCM) is a major cause of heart failure that disproportionately affects individuals of African genetic ancestry (AFR), among whom familial clustering of disease is also more pronounced relative to those of European ancestry (EUR). However, established monogenic DCM genes, identified primarily in EUR populations, explain a smaller proportion of DCM cases in AFR populations. A recent study identified a common AFR-specific nonsense variant in *CD36* that accounts for a substantial burden of DCM in AFR. How the risk and population impact of this variant compare with those of established genetic causes of DCM is unknown.

**Objective:** To compare the contribution of a *CD36* nonsense variant to DCM risk with that of truncating variants in *TTN* and pathogenic or likely pathogenic (P/LP) variants in other established DCM genes.

**Design, Setting, and Participants:** Multicohort genetic association study including AFR and EUR participants with exome or genome sequence and DCM case status from four datasets: All of Us, Million Veteran Program, Penn Medicine Biobank, and the DCM Precision Medicine Study.

**Exposure:** Carrier status for *TTN* truncating variants, P/LP variants in 11 high confidence DCM genes, and the *CD36* nonsense variant (Y325*; 0, 1, or 2 copies).

**Main Outcomes and Measures:** Odds of DCM; prevalence of risk-variant carriers among DCM cases; and population attributable fraction (PAF) for DCM.

**Results:** Among 82,623 AFR individuals across four studies, the mean age was 53.4 years and 1,625 had DCM. *CD36* Y325* risk-allele homozygotes had 4.8-fold (95% CI, 3.1-7.3) increased odds of DCM, and *CD36* Y325* heterozygotes had 1.4-fold (95% CI, 1.2-1.7) increased odds. *TTN* truncating variants also conferred elevated risk of DCM in AFR participants (OR, 8.46; 95% CI, 5.3-12.3). Among AFR DCM cases, 2.5% were *CD36* homozygotes, second only to *TTN* truncating variants (4.3%) and exceeding all other high-confidence DCM genes combined (1.5%). In population-level analyses incorporating both heterozygous and homozygous *CD36* Y325* carriers, the population-attributable fraction for *CD36* (9.0%) surpassed that of *TTN* truncating variants (3.6%).

**Conclusions and Relevance:** An ancestry-specific *CD36* variant contributes more to DCM burden in AFR ancestry than established DCM genes, including *TTN* truncating variants, typically considered the most common genetic cause of DCM. These findings reshape the known genetic architecture of DCM in individuals of African ancestry and highlight the importance of representation in genomic research.

**Key Points:** *Question:* To what extent does a common, African ancestry-specific nonsense variant in *CD36* contribute to the genetic architecture of dilated cardiomyopathy (DCM) in individuals of African ancestry in the United States?

*Findings:* In an analysis of African ancestry individuals from multiple U.S.-based cohorts, a *CD36* nonsense variant accounted for a greater population burden of DCM than *TTN* truncating variants and pathogenic or likely pathogenic variants in other established DCM genes.

*Meaning:* A single ancestry-specific *CD36* variant substantially alters current understanding of DCM genetic architecture in individuals of African ancestry and underscores the importance of including ancestral diversity in all genetic studies.

## Introduction

A predisposition to dilated cardiomyopathy (DCM) has long been observed among individuals of African genetic ancestry (AFR), who experience a higher incidence of disease compared to those of European ancestry (EUR)^1,2^. A leading cause of heart failure and cardiac transplantation, DCM also aggregates more frequently in AFR families, suggesting a stronger contribution from inherited or other shared factors in this population^3–6^. Although pathogenic and likely pathogenic variants in more than a dozen genes have been definitively linked to DCM^5^, sequencing studies of AFR DCM cases have identified fewer pathogenic variants in established DCM genes, underscoring a disconnect between observed risk and recognized genetic causes^6^.

In a recent genome-wide association study of AFR participants, we identified a nonsense variant in *CD36* (Y325*, rs3211938, p.Tyr325Ter) associated with markedly increased risk of DCM^8^. Nearly absent among individuals of EUR ancestry, the *CD36* variant was present in approximately 16% of AFR individuals, in whom it contributed more to DCM risk than established clinical risk factors including diabetes mellitus or chronic kidney disease^8^. Unlike most established DCM genes, which influence sarcomere structure and force generation, *CD36* encodes a fatty acid transporter central to myocardial energetics, and its loss of function impairs contractility through upstream disruption of cardiac fuel utilization^9^.

In this study, we evaluated how the *CD36* Y325* variant contributes to the genetic architecture of DCM in AFR populations. Across four cohorts with substantial representation of AFR participants, we compared the contribution of *CD36* Y325* on DCM risk with that of disease-causing variants in established high evidence monogenic DCM genes, including truncating variants in *TTN,* the most common cause of genetic DCM, to clarify how ancestry-specific discoveries broaden understanding of inherited cardiovascular disease^10^.

## Methods

### Study Populations

We analyzed data from three population biobanks (the All of Us Research Program (AoU), the VA Million Veteran Program (MVP), and the Penn Medicine Biobank (PMBB)) and one clinically recruited cohort (the DCM Precision Medicine Study (DCM-PM)) with genetic information linked to electronic health records (EHR) or adjudicated clinical phenotypes. AoU is a prospective cohort study from across the United States (U.S.) linking array-based genotyping and whole-genome sequencing with EHR data under a central institutional review board (IRB)^11^. MVP is a national cohort of U.S. military veterans that links array-based genotyping and whole-genome sequencing with longitudinal VA EHR data under VA Central IRB approval^12^. PMBB enrolls patients within the Penn Medicine Health System and links array-based genotyping and whole-exome sequencing with EHR data under the University of Pennsylvania IRB approval ^13^. DCM-PM is a multi-site cohort of recruited, clinically adjudicated idiopathic DCM patients linked to whole exome sequencing, with DCM diagnoses confirmed by heart failure or transplant cardiologists^6,7,14^. All participants provided written informed consent. Additional cohort details, sequencing platforms, and quality-control procedures are provided in the Supplemental Materials.

### Genetic Exposures

The primary genetic exposure was the risk allele of the *CD36* nonsense variant Y325* (Tyr325>Term, rs3211938). *CD36* Y325* genotype was determined, with the G allele representing the risk allele for DCM. Because the *CD36* Y325* mutation is seen almost exclusively in AFR ancestry individuals (rs3211938 (T/G) - AFR MAF = 9%, EUR MAF = <0.001%), analyses of *CD36* were restricted to AFR participants.

*TTN* truncating variants (*TTNtv*) were defined as rare (minor allele frequency (MAF) < 0.1%) premature truncating variants – frameshift, stop-gain, or predicted loss-of-function splice-site variants – located within exons with high cardiac expression (percent spliced in > 90%). Annotation used the Ensembl Variant Effect Predictor 109 with LOFTEE filtering^15^.

We also evaluated pathogenic or likely pathogenic (P/LP) variants in 11 other genes with high evidence for association with DCM as curated by the Clinical Genome Resource (ClinGen) (*BAG3, DES, FLNC, LMNA, MYH7, PLN, RBM20, SCN5A, TNNC1, TNNT2, DSP*) ^5^. P/LP variants were ascertained using cohort-specific pipelines: ClinVar-based annotations for DCM-related phenotypes in AoU and MVP, genetic counselor-curated reports following ACMG-guidance in PMBB, and ACMG-guided adjudication in DCM-PM. Variants were restricted to MAF <0.1% within each ancestry group. Additional information about adjudication criteria is provided in the Supplemental Materials, and a full list of genes included in these analyses is available in Supplemental Table 1.

### Phenotypes and Covariates

DCM was defined in biobank cohorts using ICD-10 code I42.0, excluding individuals with myocardial infarction or coronary artery disease occurring before or within 30 days of cardiomyopathy diagnosis. Controls lacked DCM or heart failure. Cohort-specific censoring dates were applied (AoU: August 27^th^, 2024, MVP: January 1^st^, 2025, PMBB: January 1^st^, 2020). In DCM-PM, DCM diagnoses were clinically adjudicated by heart failure cardiologists, using established criteria such as LVEF < 50% and the exclusion of secondary causes^6,7,14^. Comorbidities including hypertension, diabetes, atrial fibrillation, aortic valve disease, hyperlipidemia, chronic obstructive pulmonary disease, coronary artery disease, chronic kidney disease, and obesity were defined using standardized EHR-based phenotyping; smoking status was obtained from questionnaires when available. Additional definitions for biobank phenotyping and clinical risk factor definitions are available in Supplemental Tables 2 and 3 respectively.

### Statistical Methods

Associations between genetic variants and DCM were evaluated using Firth penalized logistic regression within each ancestry group to account for sparse variant counts. Separate models were constructed for *TTNtv*, P/LP variants in other high evidence DCM genes, and the *CD36* Y325* variant (0, 1, or 2 copies of the risk allele). All models were adjusted for age, sex, the first 10 principal components of genetic ancestry, and cohort-specific covariates. Cohort-specific effect estimates were combined using fixed-effect meta-analysis stratified by genetic ancestry. Between-cohort heterogeneity was assessed using Cochran Q statistics. Sensitivity analyses were additionally adjusted for clinical or other genetic risk factors to evaluate for independence of associations.

To characterize the distribution of strong, monogenic contributors among DCM cases within each ancestry group, we calculated the prevalence of *TTNtv* and P/LP variants in other high evidence ClinGen-DCM genes. For *CD36*, case-level prevalence analyses focused on *CD36* Y325* homozygotes (G/G), given that this genotype represents the high-penetrance form of risk most directly comparable to established “monogenic” DCM genes.

We also estimated population-attributable fraction (PAF) to quantify the contribution of each genetic factor to DCM burden in EUR and AFR populations. For *CD36*, PAF calculations included both *CD36* Y325* heterozygous and homozygous carriers to capture the population impact of this common variant across genotype classes. Analyses were performed using R version 4.2, and fixed-effect meta-analyses were conducted using the *metafor* package^16^. Additional details of the statistical analyses are outlined in the Supplemental Materials.

## Results

### Cohort Characteristics

Characteristics for participants of EUR and AFR genetic ancestry across all contributing cohorts are summarized in Table 1. Across all population biobanks, the study comprised data from 216,440 individuals of EUR genetic ancestry and 82,118 individuals of AFR genetic ancestry, including 1,982 EUR & 1,120 AFR DCM cases. The average age of participants in this study was 58.0 (EUR: 59.7, AFR: 53.4). Clinical risk factors such as diabetes, hypertension, chronic kidney disease, coronary artery disease, obesity, and hyperlipidemia were found to have similar prevalences across EUR and AFR populations; however, atrial fibrillation was found to have a higher prevalence among EUR ancestry cohorts.

**Table 1:** Cohort Characteristics. Table 1 provides cohort characteristics for the major cohorts included in this study, namely All of Us, Million Veteran Program, Penn Medicine Biobank, and The DCM Precision Medicine Study. Cohort specific information included in this data includes N European participants, N African participants, average age of enrollment, biological sex breakdown, and the prevalence of several clinical risk factors for disease. Phenotypic constructs for these clinical risk factors are available in Supplemental Table 3.

| Clinical Characteristics | All of Us |  | Million Veteran Program |  | Penn Medicine Biobank |  | The DCM Precision Medicine Study |  |
| --- | --- | --- | --- | --- | --- | --- | --- | --- |
|  | EUR | AFR | EUR | AFR | EUR | AFR | EUR | AFR |
| **Participants** | 116,624 | 48129 | 70,836 | 23,767 | 28,980 | 10,222 | 667 | 505 |
| **N Male** | 46,115<br>(39.5%) | 21,219<br>(44.1%) | 67,269<br>(95.0%) | 21,767<br>(91.6%) | 16,070<br>(55.5%) | 3,856<br>(37.7%) | 282<br>(42.3%) | 378<br>(74.9%) |
| **Enrollment Age (SD)** | 55.5<br>(16.8) | 49.8<br>(14.5) | 67.6<br>(12.7) | 61.4<br>(11.2) | 57.3<br>(16.3) | 51.7<br>(16.1) | 55.0<br>(15.5) | 50.8<br>(13.2) |
| **Atrial Fibrillation** | 5,683<br>(4.9%) | 751<br>(1.6%) | 8,918<br>(12.6%) | 1,547<br>(6.5%) | 2,278<br>(7.9%) | 443<br>(4.3%) | 46<br>(6.9%) | 26 (5.2%) |
| **Aortic Valve Disease** | 2,787<br>(2.3%) | 415<br>(0.9%) | 3,622<br>(5.1%) | 779<br>(3.3%) | 3,712<br>(12.8%) | 616<br>(6.0%) | NA | NA |
| **Coronary Artery Disease** | 4,466<br>(3.8%) | 1,624<br>(3.4%) | 14,115<br>(19.9%) | 4,636<br>(19.5%) | 3,732<br>(12.9%) | 1,414<br>(13.8%) | NA | NA |
| **Chronic Obstructive Pulmonary Disease** | 7,118<br>(6.1%) | 2,875<br>(6.0%) | 15,961<br>(22.5%) | 4,669<br>(19.6%) | 3,669<br>(12.7%) | 1,551<br>(15.2%) | NA | NA |
| **Diabetes** | 11,979<br>(10.3%) | 6,286<br>(13.1%) | 29,241<br>(41.3%) | 13,019<br>(54.8%) | 5,109<br>(17.6%) | 3,567<br>(34.9%) | 136<br>(20.4%) | 155<br>(30.7%) |
| **Hyperlipidemia** | 33,715<br>(29.9%) | 7,231<br>(15.0%) | 53,610<br>(75.7%) | 16,329<br>(68.7%) | 12,900<br>(44.5%) | 4,713<br>(46.1%) | 171<br>(25.6%) | 144<br>(27.5%) |
| **Hypertension** | 36,806<br>(31.6%) | 14,796<br>(30.7%) | 57,704<br>(81.5%) | 21,075<br>(88.7%) | 14,839<br>(51.2%) | 6,767<br>(66.2%) | 278<br>(41.7%) | 342<br>(67.7%) |
| **Obesity** | 42,175<br>(36.2%) | 22,967<br>(46.7%) | 26,146<br>(36.9%) | 10,242<br>(43.1%) | 13,568<br>(46.8%) | 7,107<br>(69.5%) | 282<br>(42.3%) | 281<br>(55.6%) |
| **Smoking Status [N ever smoker]** | 48,829<br>(41.9%) | 22,493<br>(47.7%) | 51,636<br>(72.9%) | 18,077<br>(76.1%) | 13,637<br>(47.1%) | 5,019<br>(49.1%) | 36<br>(5.4%) | 34 (6.8%) |
| **Dilated Cardiomyopathy** | 485<br>(0.4%) | 226<br>(0.5%) | 964<br>(1.8%) | 606<br>(3.4%) | 533<br>(2.2%) | 288<br>(3.5%) | 667<br>(100.0%) | 505<br>(100.0%) |
| **Heart Failure** | 7,156<br>(6.4%) | 3,146<br>(6.5%) | 4,246<br>(7.5%) | 1,904<br>(10.1%) | 5,152<br>(17.8%) | 2,222<br>(21.7%) | NA | NA |

### Population Frequency of P/LP DCM and *CD36* Y325* Variants

We first quantified population frequencies of P/LP variants in high evidence DCM variants and *CD36* Y325*. *TTN*tv were observed at similar frequencies in EUR and AFR study participants (EUR: 0.53%, AFR: 0.54%). In contrast, P/LP variants in other high evidence DCM genes were more frequent in EUR than AFR participants (EUR: 0.20%; AFR: 0.07%). Among AFR participants, *CD36* Y325* heterozygotes comprised 16.3%, while *CD36* Y325* homozygotes comprised 0.82%. The prevalence of risk gene carriers in each individual cohort is available in Supplemental Table 4.

### Genetic Risk of DCM by Variant Class and Ancestry

We next estimated the magnitude of DCM risk associated with each genetic contributor within EUR and AFR populations. *TTN*tv carrier status was associated with increased odds of DCM in both EUR and AFR populations (EUR: OR, 13.6 [95% CI: 10.8-17.3, P<0.001]; AFR: 8.5 [95% CI: 5.5-13.0, *P* <0.001]; Figure 1). These associations remained consistent when examined across several population biobanks (Supplemental Figure 1).

**Figure 1:**
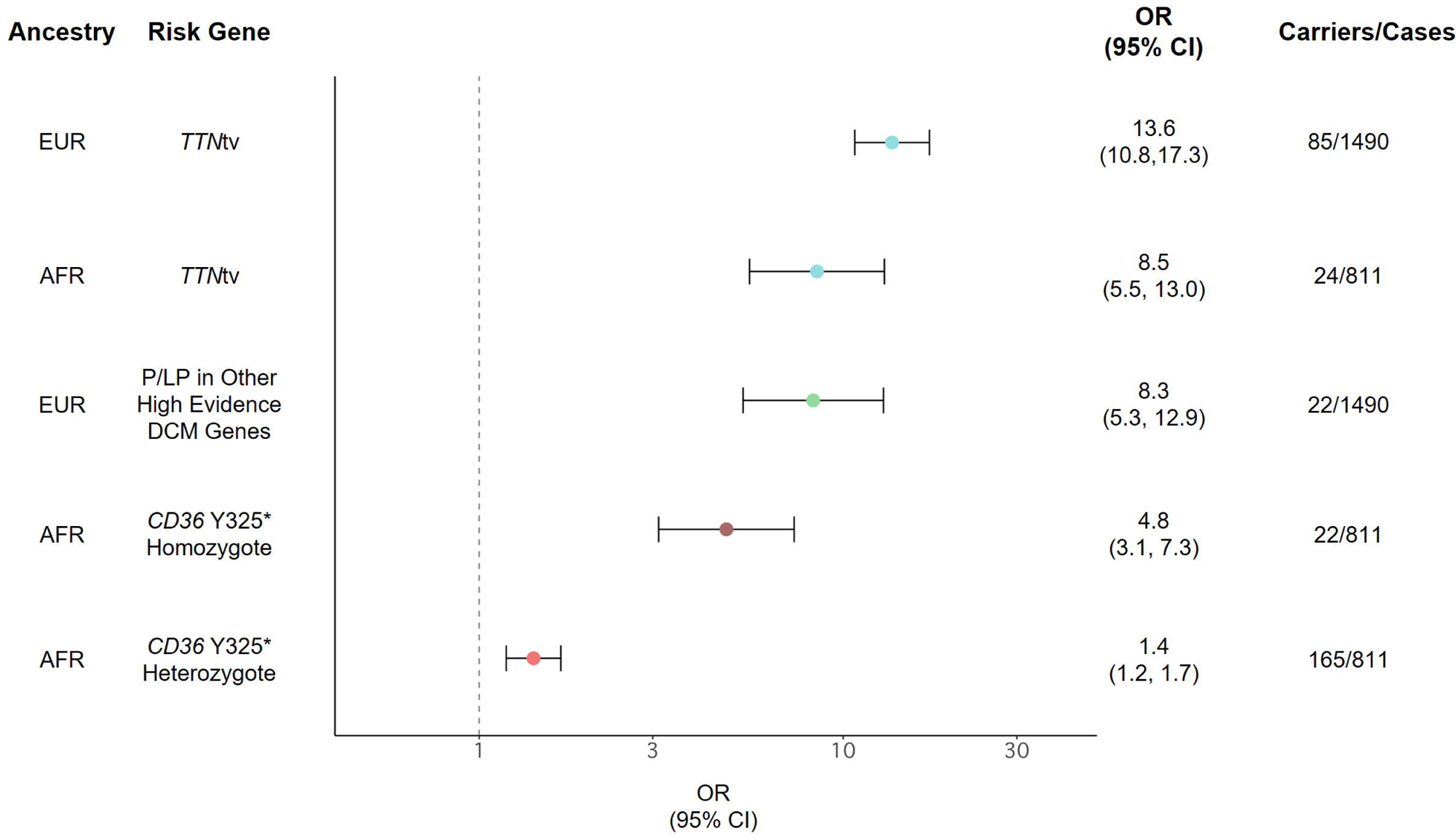
Genetic Risk of Dilated Cardiomyopathy Across European and African Populations. Figure shows the fixed effect meta-analyzed dilated cardiomyopathy (DCM) risk conferred by *TTN* truncating variants (*TTN*tv), pathogenic/likely pathogenic (P/LP) variants in other high evidence ClinGen-DCM genes, and *CD36* Y325* risk allele carriers in European (EUR) and African (AFR) ancestry populations and uses several biobanks including All of Us, Million Veteran Program, and Penn Medicine Biobank. We lacked sufficient carriers of the P/LP variants among AFR DCM cases and *CD36* Y325* carriers in EUR DCM cases to interrogate their effects.

In EUR ancestry, carriers of P/LP variants in other high evidence DCM genes also had increased risk of DCM (OR, 8.3 [95% CI: 5.3-12.9], *P*<0.001; Figure 1). In AFR participants, the number of DCM cases who harbor P/LP variants in other high evidence DCM genes was limited, precluding precise ancestry-specific risk estimation.

In AFR participants, *CD36* Y325* genotype was associated with increased DCM risk in a dose-dependent manner: heterozygotes had 1.4-fold odds of DCM (95% CI: 1.2-1.7, *P:* <0.001) and homozygotes had 4.8-fold odds (95% CI: 3.1-7.3, *P:* <0.001; Figure 1). Associations were consistent across cohorts and persisted after adjustment for clinical risk factors when examined in AoU (Supplemental Figure 2).

### Prevalence of Genetic Contributors Among DCM Cases

In an ancestry-stratified manner, we then compared *CD36* with high evidence monogenic contributors of DCM by examining only DCM cases to quantify the proportion carrying (i) a *TTN*tv, (ii) a P/LP variant in another high evidence DCM gene, or (iii) two copies of *CD36* Y325*. Prior risk modelling demonstrated that *CD36* Y325* homozygotes carried a risk profile similar in magnitude to monogenic contributors of DCM. Among 2,157 EUR DCM cases, 8.7% carried a *TTNtv* and 4.5% carried a P/LP variant in another high evidence DCM gene (Figure 2A). The distribution of genetic contributors was similar regardless of recruitment strategy (biobank vs clinical cohort) (Supplemental Figure 3).

**Figure 2:**
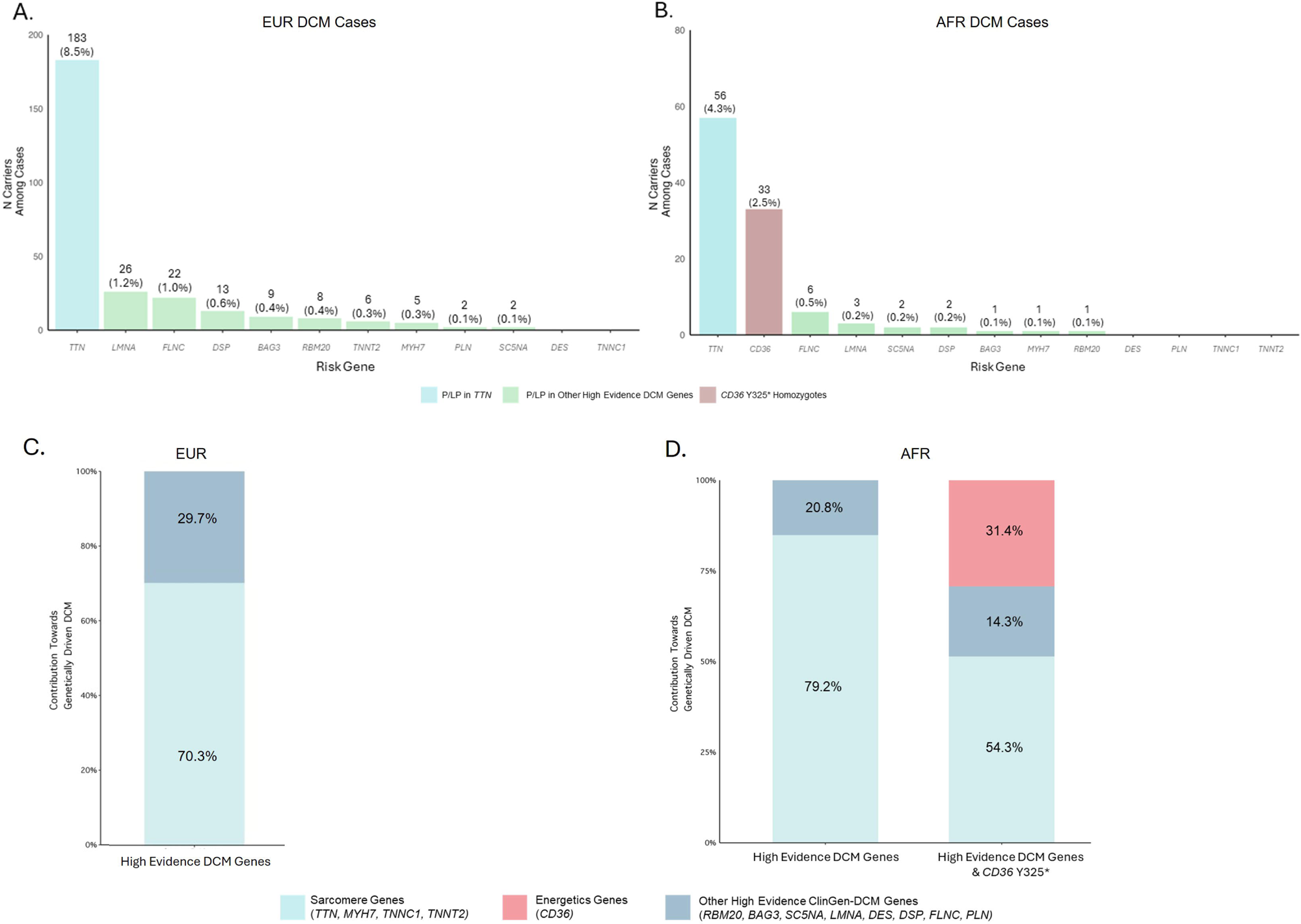
Prevalence of Dilated Cardiomyopathy (DCM) Genes in European and African Cases. Panels A & B shows the prevalence of pathogenic/likely pathogenic (P/LP) variant carriers in high evidence ClinGen-DCM genes and *CD36* Y325* risk-allele homozygote carriers among European (A) and African (B) DCM cases. Panels C & D shows the contribution of the same genes stratified by the implicated pathways of their final protein product. Panel C shows results for EUR [N(high evidence)=278] and Panel D for AFR DCM cases [N(high evidence)=78, N(high evidence + *CD36* Y325*)=106]. This figure incorporates data from several cohorts including All of Us, Penn Medicine Biobank, The DCM Precision Medicine Study, and the Million Veteran Program.

Among 1,316 AFR DCM cases, 4.3% carried at least one *TTNtv*, and 1.5% carried a P/LP variant in another high evidence DCM gene. 2.5% were homozygous carriers of *CD36* Y325* (Figure 2A). Similar to observations in EUR, the rank-order of risk gene prevalences was similar when stratified by recruitment strategy (Supplemental Figure 3). The complete prevalence of risk gene carriers among DCM cases is available in Supplemental Table 5.

### Genetic Architecture of DCM in EUR and AFR populations

We next evaluated the distribution of high-impact genetic contributors among DCM cases with an established monogenic contributor (a P/LP variant in any high evidence DCM gene, including *TTN*) within each ancestry group and then repeated this analysis with the addition of *CD36* Y325* homozygotes. Genes were further grouped into their mechanistic pathways, as previously described^5^.

When examining DCM cases arising from P/LP variants in all high evidence ClinGen-DCM genes, in EUR (N=276) 70.3% carried a P/LP variant in those implicated in sarcomere function (*TTN, MYH7, TNNC1, TNNT2)* (Figure 2B). Due to its extremely rare frequency in EUR, none carried *CD36* Y325*. Among AFR DCM cases, 79.2% carried a P/LP variant in genes implicated in sarcomere function (*TTN, MYH7, TNNC1, TNNT2)* (Figure 2B). When *CD36* Y325* homozygotes were added, the proportion of AFR cases arising from a genetic contributor increased from 14.3% (N=72) to 35.8% (N=105). Within this expanded subset, 54.3% carried P/LP variants in a sarcomere gene, 31.4% were *CD36* Y325* homozygotes (involving a myocardial energetic pathway), and 14.3% were in genes with other functions (Figure 2B). Co-occurrence of *CD36* Y325* homozygotes with *TTNtv* or P/LP variants in other high evidence DCM genes was uncommon (Supplemental Figure 4).

### Population-Level Contribution of Genetic Factors in EUR and AFR Ancestry

While only *CD36* Y325* homozygotes were considered in the prior case-only analyses, *CD36* Y325* heterozygotes were also associated with significantly increased odds of DCM. 28.8% of AFR DCM cases carried at least one copy of the *CD36* Y325* risk allele, therefore we examined the population-level risk posed by all carriers of this variant using population-attributable fraction and compared it to *TTN*.

We first established that, among EUR ancestry individuals, the PAF for *TTN*tv (6.0%) far exceeded that for P/LP mutations in all other high evidence DCM risk genes combined (0.6%) [Figure 3a], which was consistent across individual cohorts [Supplemental Figure 5]. By comparison, in AFR ancestry individuals, the *CD36* Y325* variant had a PAF of 9.0%, with contributions from heterozygotes (6.2%) and homozygotes (2.8%), while *TTN*tv had a PAF of 3.7% (Figure 3b). The higher PAF of *CD36* Y325* relative to *TTN*tv remained consistent when examined in individual population biobanks [Supplemental Figure 5]. Sparse counts precluded precise PAF estimation for other high-evidence DCM genes in AFR individuals; nonetheless, their low prevalence suggests a limited population-attributable contribution in these cohorts.

**Figure 3:**
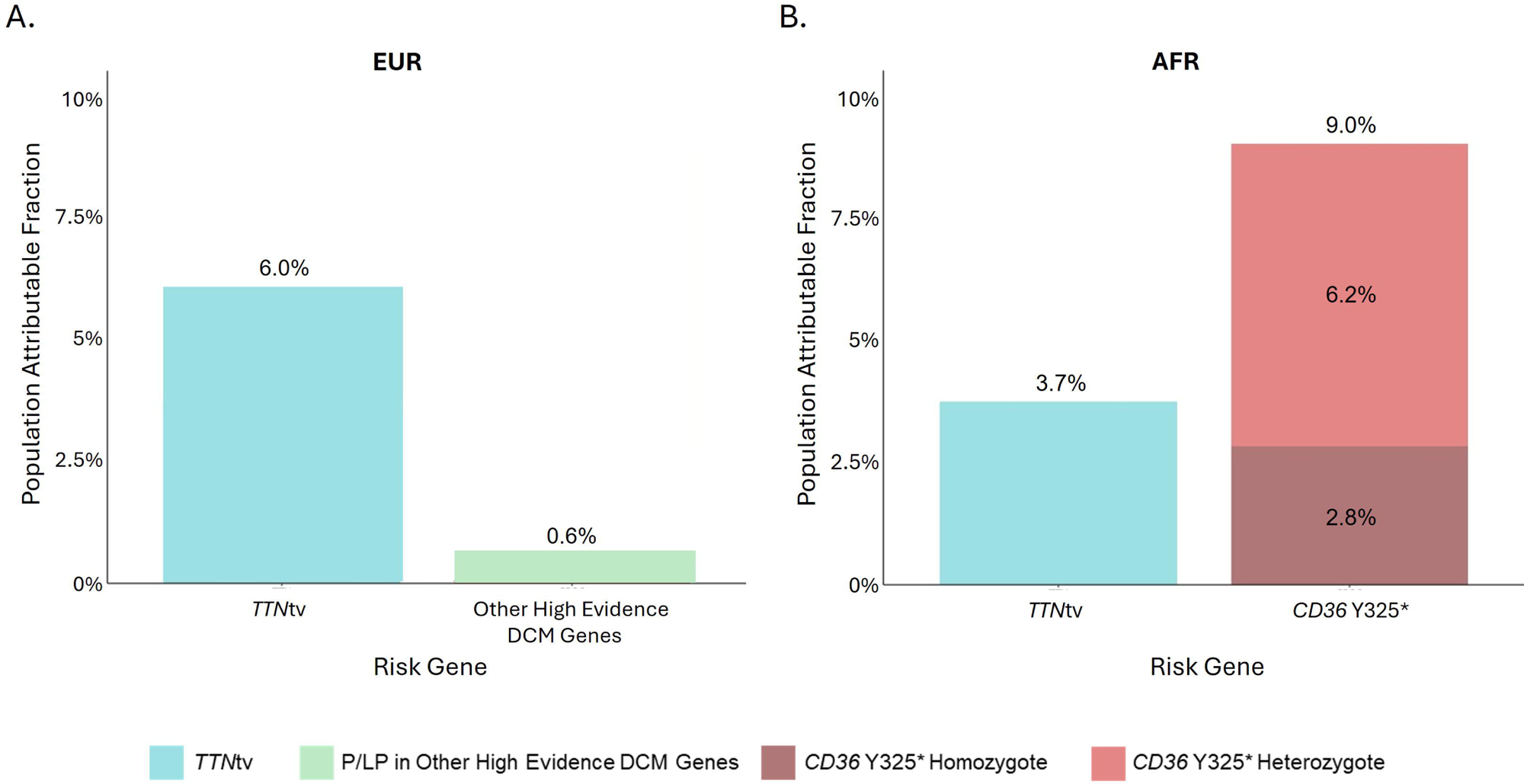
Population Attributable Fraction of DCM Risk Genes in European and African Populations. Figure shows the population attributable risk of *TTN* truncating variants (*TTN*tv) and other high evidence ClinGen-DCM genes in European ancestry (EUR) participants (Panel A) and *TTN*tv and *CD36* Y325* in African ancestry (AFR) populations (Panel B). This figure incorporates fixed effect modeling using risk estimates derived from several population biobanks including All of Us, Million Veteran Program, and Penn Medicine Biobank and also uses the overall prevalence of these genetic factors across these populations.

## Discussion

In this multi-cohort genetic association study, the African ancestry-specific *CD36* nonsense variant Y325* was strongly associated with DCM and accounted for a substantial share of DCM genetic susceptibility in individuals of African descent. In population-attributable fraction analyses, the estimated contribution of the *CD36* variant exceeded that of *TTN* truncating variants and other established monogenic DCM genes, reframing contemporary estimates of inherited DCM burden in individuals of AFR ancestry.

Individuals of African ancestry experience approximately twice the incidence of heart failure and DCM compared with individuals of European ancestry, with earlier disease onset and worse outcomes^2,17,18^. Differences in clinical risk factors and structural inequities in care may contribute to this disparity but do not fully account for the outsized predilection for DCM historically observed in AFR populations. In a recent genome-wide association study, we identified an AFR-specific nonsense variant in *CD36* (Y325*) associated with DCM that accounted for approximately one-fifth of excess DCM risk in AFR populations^8^. The present study sought to extend this discovery by placing the *CD36* variant in clinical genetic context by directly comparing its association and estimated population impact with established genetic risk factors for DCM routinely evaluated in clinical practice.

Clinical genetic evaluation for DCM has traditionally centered on rare, high-impact variants in established cardiomyopathy genes^5^ with *TTN* truncating variants widely regarded as the most common genetic cause^10,19^. Notably, it has been reported that P/LP variants in high evidence DCM risk genes show a lower prevalence among AFR cases^7^; however, it has remained unclear whether this difference was due to incomplete variant adjudication or differing genetic architecture.

In the current analysis, *CD36* Y325* homozygotes, nearly 1% of AFR individuals, had approximately 5-fold higher odds of DCM, a magnitude typically attributed to canonical monogenic contributors. Among AFR DCM cases, *CD36* Y325* homozygotes were second only to *TTN* truncating variants in case prevalence and exceeded the combined prevalence of variants in the next 11 high evidence ClinGen-DCM genes. Co-occurrence of P/LP variants in high evidence DCM genes in C*D36* Y325* homozygotes was uncommon, and inclusion of *CD36* Y325* homozygotes increased the proportion of AFR DCM cases with an identifiable monogenic cause by more than one-third. The findings therefore support *CD36* Y325* homozygosity as a distinct, prevalent, and high-impact genetic contributor to DCM in AFR populations that warrants consideration alongside established high evidence DCM genes.

These data underscore the necessity of comprehensive studies of populations beyond just European ancestry to fully understand DCM genetics and the interplay of common and rare variant effects. Beyond the high-impact homozygous state, *CD36* Y325* heterozygotes, 16% of AFR individuals, were also associated with increased odds of DCM at an effect size (OR ∼1.4) seldom observed for a common variant in complex disease^20,21^. When both heterozygous and homozygous genotypes were incorporated into population-attributable fraction analyses, *CD36* Y325* accounted for an estimated 9% of DCM burden in AFR populations, exceeding corresponding estimates for *TTN* truncating variants and other established DCM genes. Together, these observations inform the emphasis on *TTN* as the primary genetic contributor to DCM – an inference derived largely from European ancestry cohorts – by demonstrating that, in AFR individuals, a single variant in *CD36* represents the largest single-gene contributor to DCM, an observation essential to full insight into DCM risk for individuals of African ancestry.

The large population impact of the *CD36* Y325* variant on DCM risk in AFR populations is notable given the biological mechanism it implicates. Whereas most genes emphasized in contemporary DCM panels map to sarcomere structure, cytoskeletal integrity, or nuclear-envelope biology, CD36 directly influences myocardial fatty acid uptake and substrate utilization^22–25^. The prominence of *CD36* Y325* in AFR DCM therefore shifts the distribution of genetically implicated pathways toward myocardial energetics and metabolism, highlighting energetic mechanisms as an important component of disease biology^26^. More broadly, these data show that ancestry-specific genetic variation can reshape both gene-level estimates of inherited DCM burden as well as the biological pathways prioritized in DCM pathogenesis.

These observations provide compelling evidence that a clinical genetic evaluation of a patient with DCM of African ancestry would be informed by determining *CD36* Y325* genotype to identify homozygous carriers. Furthermore, identifying heterozygotes and defining more clearly their risk of DCM in future studies will be important to guide appropriate clinical interpretation and implementation. Finally, as genotype-informed precision care evolves^27^, the outsized contribution of the *CD36* Y325* in AFR populations underscores the need for precision therapy frameworks that also incorporate energetic mechanisms.

## Limitations

Several limitations should be acknowledged. First, DCM ascertainment in population biobanks relied largely on diagnosis code-based definitions, which may introduce misclassification. Mitigating this limitation is the concurrence of observations from population biobanks and the recruited DCM-PM study. Second, P/LP variants in high evidence DCM genes in the population biobanks were identified using available annotation resources (including ClinVar and *in-silico* prediction) and may be incomplete, particularly in non-European populations where variant classification is less mature. Third, cohorts differed in recruitment strategy and variant curation workflows, which may affect comparability of prevalence and effect estimates despite generally concordant patterns across datasets. Fourth, population-attributable estimates are model-based and focused on *CD36* Y325* (and selected monogenic comparators); these analyses did not capture other potentially pathogenic *CD36* alleles or common-variant/polygenic contributions and therefore should be interpreted as estimates of burden. Finally, this study focused on AFR ancestry populations from across the United States. Further work is needed to establish the burden of risk *CD36* poses on DCM among AFR populations in other countries, including among continental African populations.

## Conclusions

The African ancestry-specific *CD36* nonsense variant Y325* contributes substantially to DCM risk and burden in AFR populations from the United States, rivaling and exceeding the population-level contribution of canonical monogenic DCM genes. These findings inform the prevailing view of DCM architecture and underscore the importance of ancestry-specific discovery to inform equitable genetic evaluation and biological models of disease.

## Data Availability

All data produced in the present work are contained in the manuscript. Individual-level data for the underlying biobanks or clinical cohorts require application for access by approved researchers following study-specific mechanisms.

## Article Information

### Corresponding Authors

Krishna Aragam, MD, MS and Jennifer Huffman, PhD

### Author contributions

*Concept and design:* S. Challa, M. Levin, R. Hershberger, J. Huffman, K. Aragam

*Acquisition, analysis, or interpretation of data:* S. Challa, A. Zheng, S. Jurgens, X. Wang, S.H. Choi, J. Halford, J. Mead, P.T. Ellinor, R. Hershberger, J. Huffman, S. Damrauer, P. Tsao, K. Aragam

*Drafting of the manuscript:* S. Challa, K. Biddinger, A. Zheng, S. Abramowitz, M. Levin, R. Hershberger, J. Huffman, K. Aragam

*Critical revision of the manuscript for intellectual context:* All Co-Authors

*Statistical analysis:* S. Challa, K. Biddinger, A. Zheng, S. Abramowitz, J. Mead, D. Kinnamon, J. Huffman

*Obtaining funding:* K.-M. Chang, P. Tsao, S. Damrauer, P.T. Ellinor, R. Hershberger, K. Aragam

*Administrative, technical, or material support:* K.-M. Chang

*Supervision:* M. Levin, R. Hershberger, J. Huffman, K. Aragam

### Additional Contributions

We acknowledge the Penn Medicine BioBank (PMBB) for providing data and thank the patient-participants of Penn Medicine who consented to participate in this research program. We would also like to thank the Penn Medicine BioBank team and Regeneron Genetics Center for providing genetic variant data for analysis. The PMBB is approved under IRB protocol# 813913 and supported by Perelman School of Medicine at University of Pennsylvania, a gift from the Smilow family, and the National Center for Advancing Translational Sciences of the National Institutes of Health under CTSA award number UL1TR001878. This research has been conducted using the All of Us cohort study. We gratefully acknowledge All of Us participants for their contributions, without whom this research would not have been possible. We also thank the National Institutes of Health’s All of Us Research Program for making available the participant data examined in this study. We gratefully acknowledge the Veterans who participated in the VA Million Veteran Program.

### Additional Information

Saketh Challa, BSc^1,2,3^*; Kiran Biddinger, BSE^1,2,3,*^; Sarah Abramowitz, BA^4,5,*^; Alice Zheng, MSc^1,2^; Jonathan O. Mead, MTDA^6, 7,8^; Renae L. Judy, MS^4,9^; Sean Jurgens, MD, PhD^1,2,10^; Liam Gaziano, PhD^1,2,3^; Xin Wang, MPH^1,2^; Seung Hoan Choi, PhD^1,11^; Jenny Halford, MD^1,2^; Elizabeth Jordan, MS^6,7^,; Joseph Liu, MS^12^; VA Million Veteran Program^+^; Penn Medicine Biobank^+^; Kyong-Mi Chang, MD^9,13^; Amanda Vest, MBBS^12^; W.H. Wilson Tang, MD^12^; Philip Tsao, PhD^14,15^; Daniel D. Kinnamon, PhD^6,7,16^ ; Scott M. Damrauer, MD^4,9,13,17^; Patrick T. Ellinor, MD, PhD^1,2^; Michael Levin, MD^9,17,18^**; Ray E. Hershberger, MD^7,19^**; Jennifer E. Huffman, PhD^1,3,14,20^**; Krishna G. Aragam MD, MS^1,3,12^**

