## Supplemental Materials, Figures, and Tables for "A Common *CD36* Variant and the Genetic Landscape of Dilated Cardiomyopathy in Individuals of African Ancestry"


**Supplemental Acknowledgements** **2**

**Supplemental Methods** **4**

**Supplemental Figures** **9**

**Supplemental Tables** **15**

**References** **20**

### Supplemental Acknowledgements

#### A. Million Veteran Program


MVP Program Office

- Sumitra Muralidhar, Ph.D., Program Director
  US Department of Veterans Affairs, 810 Vermont Avenue NW, Washington, DC 20420
- Jennifer Moser, Ph.D., Associate Director, Scientific Programs
  US Department of Veterans Affairs, 810 Vermont Avenue NW, Washington, DC 20420
- Jennifer E. Deen, B.S., Associate Director, Cohort & Public Relations
  US Department of Veterans Affairs, 810 Vermont Avenue NW, Washington, DC 20420

MVP Steering Committee

- Co-Chair: Philip S. Tsao, Ph.D.
  VA Palo Alto Health Care System, 3801 Miranda Avenue, Palo Alto, CA 94304
- Co-Chair: Sumitra Muralidhar, Ph.D.
  US Department of Veterans Affairs, 810 Vermont Avenue NW, Washington, DC 20420
- J. Michael Gaziano, M.D., M.P.H.
  VA Boston Healthcare System, 150 S. Huntington Avenue, Boston, MA 02130
- Adriana Hung, M.D., M.P.H.
  VA Tennessee Valley Healthcare System, 1310 24th Ave. South, Nashville, TN 37212
- Dave Oslin, M.D.
  Philadelphia VA Medical Center, 3900 Woodland Avenue, Philadelphia, PA 19104
- Deepak Voora, MD
  Durham VA Medical Center, 508 Fulton Street, Durham, NC 27705

MVP Co-Principal Investigators

- J. Michael Gaziano, M.D., M.P.H.
  VA Boston Healthcare System, 150 S. Huntington Avenue, Boston, MA 02130
- Philip S. Tsao, Ph.D.
  VA Palo Alto Health Care System, 3801 Miranda Avenue, Palo Alto, CA 94304

MVP Core Operations

- Jessica V. Brewer, M.P.H., Director, MVP Cohort Operations
  VA Boston Healthcare System, 150 S. Huntington Avenue, Boston, MA 02130
- Mary T. Brophy M.D., M.P.H., Director, VA Central Biorepository
  VA Boston Healthcare System, 150 S. Huntington Avenue, Boston, MA 02130
- Kelly Cho, M.P.H, Ph.D., Director, MVP Phenomics
  VA Boston Healthcare System, 150 S. Huntington Avenue, Boston, MA 02130
- Lori Churby, B.S., Director, MVP Regulatory Affairs
  VA Palo Alto Health Care System, 3801 Miranda Avenue, Palo Alto, CA 94304
- Jacob T. Kean, Ph.D., Acting Director, VA Informatics and Computing Infrastructure (VINCI)
  VA Salt Lake City Health Care System, 500 Foothill Drive, Salt Lake City, UT 84148
- Saiju Pyarajan Ph.D., Director, Data and Computational Sciences
  VA Boston Healthcare System, 150 S. Huntington Avenue, Boston, MA 02130
- Robert Ringer, Pharm.D., Director, VA Albuquerque Central Biorepository
  New Mexico VA Health Care System, 1501 San Pedro Drive SE, Albuquerque, NM 87108
- Luis E. Selva, Ph.D., Director, MVP Biorepository Coordination
  VA Boston Healthcare System, 150 S. Huntington Avenue, Boston, MA 02130
- Shahpoor (Alex) Shayan, M.S., Director, MVP PRE Informatics

VA Boston Healthcare System, 150 S. Huntington Avenue, Boston, MA 02130
- Brady Stephens, M.S., Principal Investigator, MVP Information Center
Canandaigua VA Medical Center, 400 Fort Hill Avenue, Canandaigua, NY 14424
- Stacey B. Whitbourne, Ph.D., Director, MVP Cohort Development and Management
VA Boston Healthcare System, 150 S. Huntington Avenue, Boston, MA 02130

#### B. Penn Medicine Biobank

PMBB Leadership Team
Daniel J. Rader, M.D., Marylyn D. Ritchie, Ph.D.
**Contribution**: All authors contributed to securing funding, study design and oversight. All authors reviewed the final version of the manuscript.

Patient Recruitment and Regulatory Oversight
JoEllen Weaver, Nawar Naseer, Ph.D., M.P.H., Giorgio Sirugo, M.D., P.h.D., Afiya Poindexter, Jenna Dever, Aidan Harvey, Sydney Linn, Naman Srivastava
**Contributions**: JW manages patient recruitment and regulatory oversight of study. NN manages participant engagement, assists with regulatory oversight, and researcher access. GS assists with researcher access. AP, JD, AH, SL, and NS perform recruitment and enrollment of study participants.

Lab Operations
JoEllen Weaver, Meghan Livingstone, Fred Vadivieso, Stephanie DerOhannessian, Teo Tran, Julia Stephanowski, Salma Santos, Ned Haubein, P.h.D., Joseph Dunn
**Contribution**: JW, ML, FV, SD conduct oversight of lab operations. ML, FV, AK, SD, TT, JS, SS perform sample processing. NH, JD are responsible for sample tracking and the laboratory information management system.

Clinical Informatics
Anurag Verma, Ph.D., Colleen Morse Kripke, M.S. DPT, MSA, Marjorie Risman, M.S., Renae Judy, B.S., Colin Wollack, M.S.
**Contribution**: All authors contributed to the development and validation of clinical phenotypes used to identify study subjects and (when applicable) controls.

Genome Informatics
Anurag Verma Ph.D., Shefali S. Verma, Ph.D., Scott Damrauer, M.D., Yuki Bradford, M.S., Scott Dudek, M.S., Theodore Drivas, M.D., Ph.D., Zachary Rodriguez, Ph.D,
**Contribution**: AV, SSV, and SD are responsible for the analysis, design, and infrastructure needed to quality control genotype and exome data. YB performs the analysis. TD and AV provides variant and gene annotations and their functional interpretation of variants.

For PMBB, please use:
For Regeneron, please use:

### Supplemental Methods

#### A. Study Populations

##### All of Us

The All of Us Research Program (AoU) is a cohort study from the United States that is currently recruiting participants across the country. AoU has whole-genome sequencing data from 245,388 individuals and detailed phenotypic data from electronic health records. The inclusion of AoU data in this study was approved under a data agreement between Massachusetts General Hospital and AoU. Study participants provided written informed consent, and genetic data methods and quality control within AoU has previously been reported[1]. Genotyping and quality control in All of Us have been reported previously (https://support.researchallofus.org/hc/en-us/articles/29475228181908-How-the-All-of-Us-Genomic-data-are-organized) and details for this process are briefly summarized below.

##### Million Veteran Program

The Veterans Affairs (VA) Million Veteran Program (MVP) started recruiting US military Veterans from 63 VA facilities across the United States in 2011. Veterans aged 18 years and older are recruited into MVP where participants are linked to VA electronic health records (EHR), complete a questionnaire, and submit a blood sample at enrollment. The EHR includes information on inpatient International Classification of Disease (ICD) diagnosis codes, Current Procedural Terminology (CPT) procedure codes, and clinical laboratory measurements[2]. Genotyping and quality control in MVP have been reported previously[3,4] and details for whole genome sequencing are briefly summarized below.

##### Penn Medicine Biobank

The Penn Medicine Biobank (PMBB) recruits from the Penn Medicine health system and links individuals' data to the EHR, which includes elements like those found in MVP. This EHR dataset was combined with 39,202 PMBB volunteers with whole-exome sequencing data to derive disease status, variant carrier status, age, and biological sex. Genotyping and quality control in PMBB have been previously reported[5] and sequencing methods are summarized in detail below.

##### The DCM Precision Medicine Study

The DCM Precision Medicine Study (DCM-PM) is a study of recruited DCM patients from sites across the United States. Patients were recruited into the study upon presentation of DCM verified by a heart failure or heart transplant cardiologist, and all analyses were restricted to patients over the age of 15. DCM verification was based on a left ventricular ejection fraction (LVEF) less than 50% and left ventricular enlargement, with other causes excluded. DCM diagnoses were validated by available magnetic resonance imaging data[6]. All participants gave written informed consent, and the institutional review boards at The Ohio State University and other clinical sites provided initial approval, followed by approval from a single institutional review board at the University of Pennsylvania. Details regarding study design and recruitment have previously been described[7–9].

#### B. Sequencing and Quality Control

##### All of Us

Each Genome Center performed quality control of specimens obtained from the All of Us Biobank. Sample preparation, normalization, and DNA library construction have all been previously reported[1]. Sample preparation, normalization, and DNA library construction have all been previously reported, and after undergoing this process samples underwent whole genome sequencing (WGS). This process consisted of initial per-sample QC that included fingerprint concordance (array vs. WGS data), sex concordance (self-report vs. genetically determined), cross-individual contamination rate and coverage aimed at detecting major errors such as sample swaps or contamination. Individuals who were flagged as failing any of these QC metrics were excluded from all subsequent releases. The whole genome sequencing variants were jointly called, and additional sample QC procedures such as hard threshold flagging and population outlier flagging were performed. Variant QC was performed following sample level QC and included hard threshold filtering (ExcessHet, QUAL score) and the Allele-Specific Variant Quality Score Recalibration (AS-VQSR).

Additional QC was performed for data that was used in the analysis of rare genetic variants. The v7 short read exome dataset with split variants was used and was restricted to genotypes that passed central QC procedures and that had a Genotype Quality value greater than 20. Filtering out variants that were identified as being monomorphic or that had call rates less than 90%. Upon removing any samples that failed QC metrics described above, 242,902 participants were left in the analysis cohort. Restricting to individuals with complete EHR records and to those who were found to be genetically unrelated, 195,533 samples were used in all subsequent analyses.

##### Million Veteran Program

This study used data from MVP whole genome sequencing (WGS) Data Release 2. Sequencing in MVP was performed by Personalis using Illumina NovaSeq6000 technology, targeting a 30x coverage. FastQC (v0.11.4) (https://www.bioinformatics.babraham.ac.uk/projects/fastqc/) was used to verify individual genome sequencing read quality, Samtools[10,11] (v0.1.19) for read alignment, and verifyBAMID[12] to measure contamination rate. Individual genome variant calling was completed using GATK[13–15] pipeline and genomes aggregated using VDS Combiner[16]. All WGS data processing and quality control was performed in Google Cloud Platform. The final dataset for analysis comprised of 104,923 participants and 663,351,127 variants. Average read depth was 27.96, average genotype quality was 73.63, and the mean sample call rate was 0.99 across all chromosomes.

##### Penn Medicine Biobank

Whole exome sequencing (WES) was performed on approximately 44,000 individuals through a collaboration with the Regeneron Genetics Center. Sample level QC was applied and sample sex errors, high rates of contamination, samples with low sequence coverage, and genetically identified sample duplicates were flagged and subsequently excluded.

##### The DCM Precision Medicine Study

Research exome sequencing and array-based genotyping of probands and affected family members was conducted at the University of Washington Northwest Genomics Center and data processed as described previously. Exome sequencing used the Roche/NimbleGen SeqCap EZ v2.0 capture, and array-based genotypes were obtained using the Illumina Global Screening Array v1.0 and v2.0. Rigorous sequencing quality control along with variant-level and sample-level quality control procedures for array-based genotypes that accounted for the diverse ancestry of study subjects are detailed elsewhere[9,17].

#### C. Principal Component Assignment

##### All of Us

All of Us applied the analysis pipeline from the gnomAD v3.1 release to calculate the principal components and assign ancestry labels. AoU first identified 150,229 highly-quality sites that were accurately called in both the Human Genome Diversity Project (HGDP) and in the 1000 Genomes (1000G) dataset. These high-quality sites were defined as being autosomal bi-allelic single nucleotide variants (SNVs) with a minor allele frequency greater than 0.1% and a call rate greater than 99%. These variants were then LD-pruned with a cut off of $r^2$ greater than 0.1. AoU centrally calculated the first 16 PCs in the training set and used the hwe_normalized_pca() HAIL function with high quality SNVs to project the AoU samples into PC space, thus calculating the first 16 PCs. The first 16 PCs were also used to assign genetically inferred ancestry labels as described below. Further details regarding the way PCs were calculated have previously been described[1].

**Million Veteran Program**
Principal components were defined in MVP through the combination of 1000G data and the EIGENSOFT[18] package. The 1000 Genome Project Dataset was merged with the MVP dataset and markers with MAF less than 1% and any samples constituting related pairs were removed. MVP samples were projected onto the 1000 Genomes Project dataset with EIGENSOFT (v 6.0.1).

**Penn Medicine Biobank**
Principal components were defined in Penn Medicine Biobank in an analgous fashion to the Million Veteran Program. Samples and variants that passed QC metrics were projected with EIGENSOFT (v 7.2.0) to generate PCs that adjust for population substructure. Further details regarding this process have previously been reported[5].

**The DCM Precision Medicine Study**
Ancestry principal component scores were obtained by projecting DCM Precision Medicine Study participants onto the PC space learned from the 1000 Genomes Phase 3 integrated callset with the Online Augmentation, Decomposition, and Procrustes approach implemented in the bed_project SelfPCA function of the bigsnpr[19] R package, as previously reported[9].

#### D. Ancestry Assignments

**All of Us**
The process of assigning genetically inferred ancestry has previously been described within All of Us[1]. Briefly, a random forest classifier was trained on the HGDP and 1000G samples with known ancestry labels using 16 PCs. Ancestry categories were assigned based on gnomAD, HGDP, and 1000G and included African, Latino/Native American/ Ad Mixed American, East Asian, Middle Easten, European, Other, and South Asian. Participants were assigned to a category based on the probability generated by the random-forest model and a cut-off of 75% was used.

**Million Veteran Program**
To estimate ancestry, we obtained a reference dataset from the 1000 Genomes Project and used the smartpca module in the EIGENSOFT[18] package to project the PC loadings from a group of unrelated individuals in the reference dataset. We merged this dataset with the MVP dataset and ran smartpca to project the PCA loadings from the reference dataset. We trained a random forest classifier using continental ancestry meta-data based on the top 10 principal components from the reference training data to define genetically inferred ancestry. We then applied this random forest to the predicted MVP PCA data and assigned ancestries to individuals with a probability greater than 50%. Those with a probability less than 50% for any particular ancestry group were excluded from the study.

**Penn Medicine Biobank**
The process of assigning genetic ancestry in Penn Medicine Biobank has previously been described[5]. Briefly, the smartpca tool and the 1KGP dataset was used along with a K-means clustering approach to assign samples to the 1KGP super populations using genetic ancestry labels.

**The DCM Precision Medicine Study**
The process of assigning genetically inferred ancestry has been described previously[9] but is summarized here. Global genomic ancestry proportions were inferred from Illumina Global Screening Array genotypes using ADMIXTURE (version 1.3.0) with the 1000 Genomes Phase 3 integrated call set as the reference.

An individual's ancestry group was defined as the inferred continental ancestry group (African, East Asian, European, Native American, or South Asian) accounting for the highest proportion of that individual's genomic ancestry.

#### E. Variant Classification

##### High Evidence DCM Risk Genes

12 genes (*BAG3, DES, FLNC, LMNA, MYH7, PLN, RBM20, SC5NA, TNNC1, TNNT2, DSP, TTN*) have previously been shown to comprise the majority of genetically driven DCM risk and are collectively referred to as "ClinGen-DCM"[20]. Among these genes, truncating variants in the gene titin (*TTN*) are known to contribute towards the majority of DCM risk[18].

In this analysis, we examined two groups of variant carriers (i) *TTN* truncating variants (*TTN*tv) and (ii) pathogenic or likely pathogenic (P/LP) variants in other high evidence ClinGen-DCM genes. Variant classifications differed slightly between population biobanks and the clinical DCM-PM study and are further described below.

*i) Population Biobanks*

Within population biobanks, variant curation was completed for all variants found in each ClinGen-DCM gene using the genomic coordinates for each gene as specified in ENSEMBL. Carrier status was defined for only EUR and AFR participants and variants were further filtered to those only within a MAF<0.1% within each cohort. P/LP variants in *TTN* were identified based on whether they caused a premature truncation (stop-gain, frameshift, splice-site) within the constituent cardiac exons of the protein. These variants were identified using the software annotation tools VEP109 and its LOFTEE (https://github.com/konradjk/loftee) plugin, and individuals were termed to be carriers based on whether they harbored one of these variants (*TTN*tv).

In AoU and MVP, P/LP variants in other high evidence ClinGen-DCM genes were identified by selecting variants within the ClinVar database that are implicated in DCM development through endophenotypes[21]. In the PMBB, P/LP variants in other high evidence ClinGen-DCM genes were selected using genetic counsellor reports. Carriers were defined as individuals who carry at least one of these variants in any of the 11 genes. We employed a similar approach for selecting P/LP variants in DCM-Panel genes for further sensitivity analyses.

*ii) DCM-PM Study*

Details regarding variant adjudication in the DCM-PM study have been previously described[9,17]. Briefly, rare protein-altering variants in 36 DCM genes were adjudicated using American College of Medical Genetics/Association of Molecular Pathologists (ACMG/AMP) and the Clinical Genome Resource (ClinGen)-based criteria tailored to DCM. After automated filtering, variants not computationally adjudicated as benign, likely benign, unlikely to impact protein function, or low quality were manually reviewed and certified for a final classification and confirmed with Sanger sequencing (pathogenic, likely pathogenic, and uncertain significance only).

##### *CD36* Carrier Status

rs3211938 (Y325*) in *CD36* is an AFR-specific variant which has previously been identified as a DCM risk variant with the G allele conferring increased risk[22]. *CD36 Y325** carrier status was defined in AFR populations only, as it is a common variant. Three carrier status categories (0/1/2) were created based on the number of risk alleles carried, zero for wildtype homozygotes (T/T), one for heterozygotes (T/G), and two for risk homozygotes (G/G).

#### F. Statistical Analyses

Major statistical analyses were performed in *R* (version 4.2). Firth's logistic regression was used to model the DCM risk posed by risk gene carriers in a cohort and ancestry-specific fashion, and these models were adjusted for age, biological sex, the first ten principal components of genetic ancestry, and cohort-specific covariates. These models were run independently for P/LP variant carriers in (i) *TTN* (carrier =1 / non-carrier=0), (ii) other high evidence ClinGen-DCM genes (carrier=1 / non-carrier = 0), and (iii) *CD36* Y325* (T/T=0, T/G=1, and G/G=2). The *R* package 'metafor' was used to construct fixed-effect models that aggregated genetic-risk estimates across individual cohorts in an ancestry-specific fashion[23]. The number of DCM cases that harbored P/LP *TTN* variants, P/LP variants in other high evidence ClinGen-DCM genes, and *CD36* Y325* risk-allele homozygotes were calculated in an ancestry specific fashion across population biobanks and in the DCM-PM. Individual cohort prevalences were aggregated in an ancestry-specific fashion, thus resulting in the prevalence of risk genes in pooled EUR and AFR DCM cases.

Genetically determined DCM cases were defined as being any case that harbored a P/LP mutation in any high evidence ClinGen-DCM risk genes (including *TTN*). Using the set of AFR and EUR cases, the contribution of ClinGen-DCM genes and *CD36* risk-allele homozygous carriers was compared in the pooled EUR and AFR DCM cases. The mutual independence of the risk conferred by P/LP mutations in *TTN*, P/LP mutations in other high evidence ClinGen-DCM genes, and *CD36* Y325* was validated by examining the joint-overlap of carriers among the set of pooled AFR DCM cases.

Population attributable fraction (PAF) is an epidemiologic metric that captures the population risk conferred by a specific factor and can be calculated by the following formula, where $f$ is the prevalence of the risk factor in the population and *RR* is the relative risk that it confers towards disease, which is well approximated by the odds ratio when the disease is rare. The population frequency of genetic factors, namely P/LP mutations in *TTN*, P/LP mutation in other high evidence ClinGen-DCM risk genes, and the frequency of *CD36* Y325* risk-allele carriers (T/G, G/G) was used alongside the meta-analyzed population biobank odds ratio estimate to derive the population attributable fraction of genetic factors towards DCM in EUR and AFR ancestry populations.

$$PAF = \frac{f * (RR-1)}{(f * (RR-1) + 1)}$$

Equation 1: Population Attributable Fraction

#### G. Supplemental Analyses

##### Independence of Genetic Risk and Clinical Risk Factors

The genetic risk posed by rs3211938 risk-allele carriers and *TTN*tv carriers was modeled in All of Us using Firth's penalized logistic regression. Carrier status was established as stated previously, and models were adjusted for age, sex, clinical factors included in Supplemental Table 5, and the first ten principal components of genetic ancestry [Supplemental Figure 2]. A full list of codes used in determining clinical risk factors is available in Supplemental Table 2.

#### Supplemental Figures

Supplemental Figure 1: Dilated Cardiomyopathy Risk Conferred by Genetic Variants in Biobank Populations.

Supplemental Figure 2: Dilated Cardiomyopathy (DCM) Risk Conferred by Genetic Variants Before and After Adjustment for Clinical Risk Factors.

Supplemental Figure 3: Prevalence of Dilated Cardiomyopathy (DCM) Genes by Ancestry and Study Recruitment Strategy.

Supplemental Figure 4: Overlap of Dilated Cardiomyopathy Risk Gene Carriers.

Supplemental Figure 5: Population Attributable Fraction of Dilated Cardiomyopathy Risk Genes Across Biobank Studies by Ancestry.

**Supplemental Figure 1: Dilated Cardiomyopathy Risk Conferred by Genetic Variants in Biobank Populations.** Panel A & B show the dilated cardiomyopathy (DCM) risk that is conferred by TTN truncating variants (TTNtv) in European (EUR) and African (AFR) ancestry populations, and includes data originating from All of Us, Penn Medicine Biobank (PMBB), and Million Veteran Program. Panel C shows the risk conferred by pathogenic or likely pathogenic (P/LP) variants in other high evidence DCM risk genes in EUR populations and includes data from All of Us, Penn Medicine Biobank, and Million Veteran Program. We lacked sufficient carriers among cases for AFR individuals. Panel D & E shows DCM risk that is conferred by *CD36* Y325* risk-allele homozygotes and heterozygotes in AFR individuals, containing data from All of Us, Penn Medicine Biobank, and Million Veteran Program. The *CD36* Y325* risk-allele was not present in EUR populations.

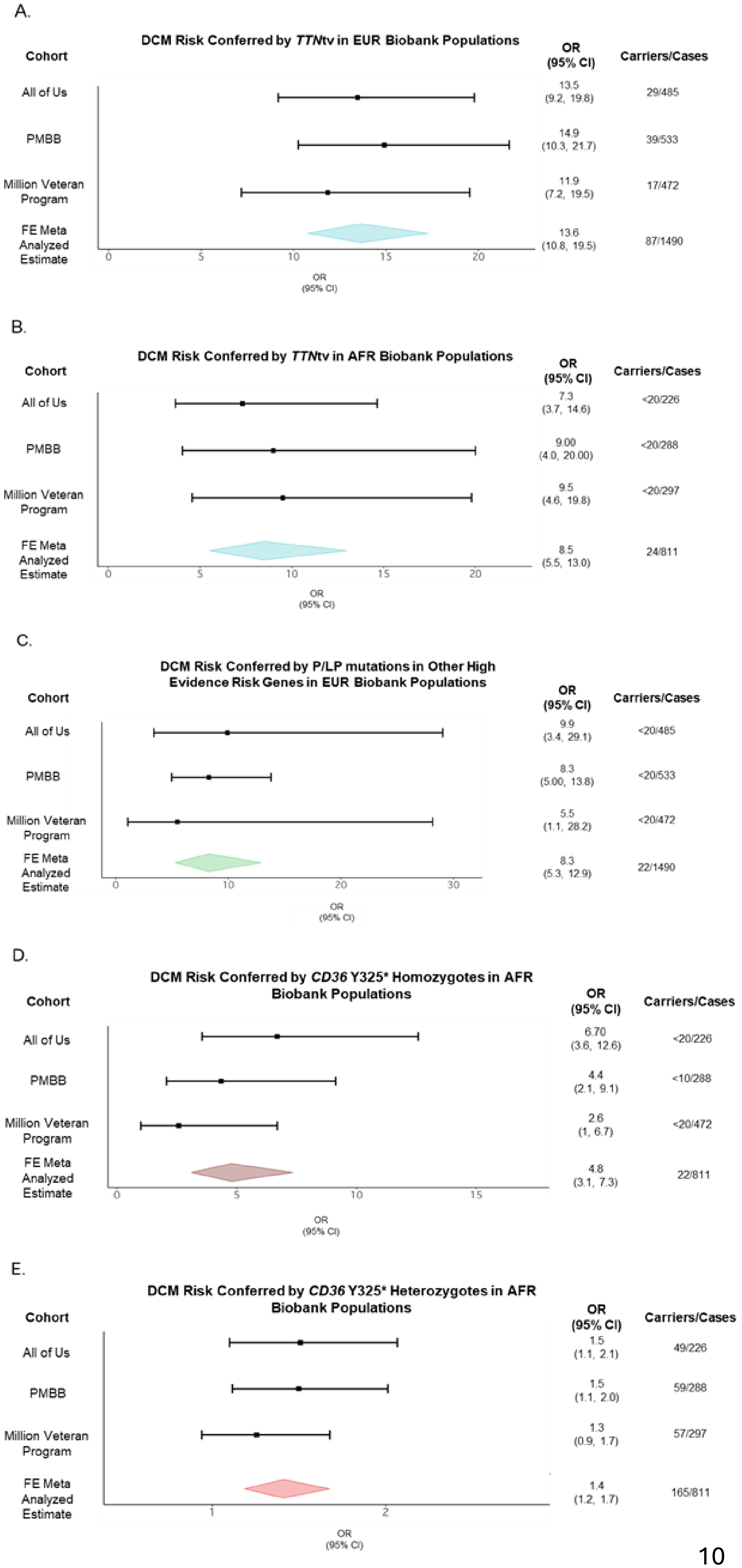

**Supplemental Figure 2: Dilated Cardiomyopathy (DCM) Risk Conferred by Genetic Variants Before and After Adjustment for Clinical Risk Factors.** This figure illustrates the DCM risk conferred by *TTN*tv and *CD36* Y325* risk-allele carriers in European and African ancestry populations in the All of Us biobank, before (Panel A) and after (Panel B) adjusting for clinical risk factors of DCM. Risk factors that were included in this analysis are atrial fibrillation, aortic valve disease, coronary artery disease, chronic obstructive pulmonary disease, diabetes, hyperlipidemia, hypertension, obesity, and smoking status. A full list of codes that are included for each clinical risk factor is available for further review in Supplemental Table 2.

A.

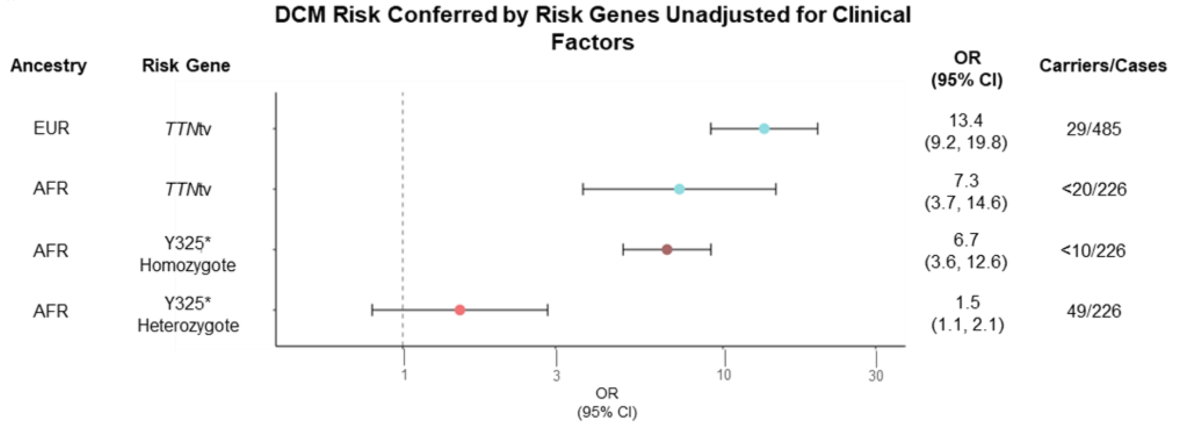


B.

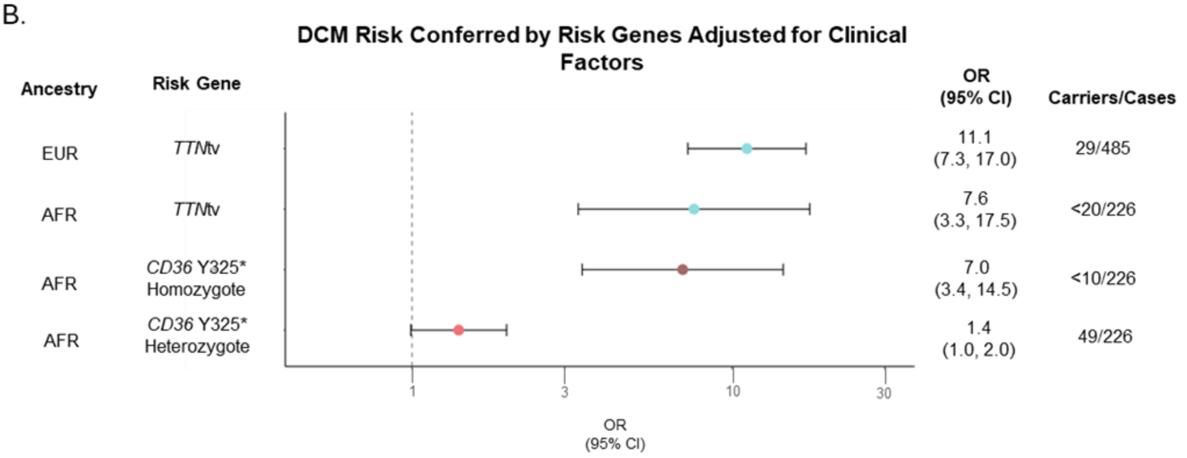

**Supplemental Figure 3: Prevalence of Dilated Cardiomyopathy (DCM) Genes by Ancestry and Study Recruitment Strategy.** Panels A & B illustrate the breakdown of EUR DCM cases when separated by recruitment strategy, population biobank (Panel A – studies = All of Us, Penn Medicine Biobank, Million Veteran Program) or recruited clinical cohort (Panel B – study = The DCM-Precision Medicine Study). Panels C & D show the same for AFR DCM cases (Panel C = population biobank, Panel D = recruited clinical cohort). Genes that were examined in this study include high-evidence ClinGen-DCM genes as well as *CD36* Y325* risk-allele homozygotes.

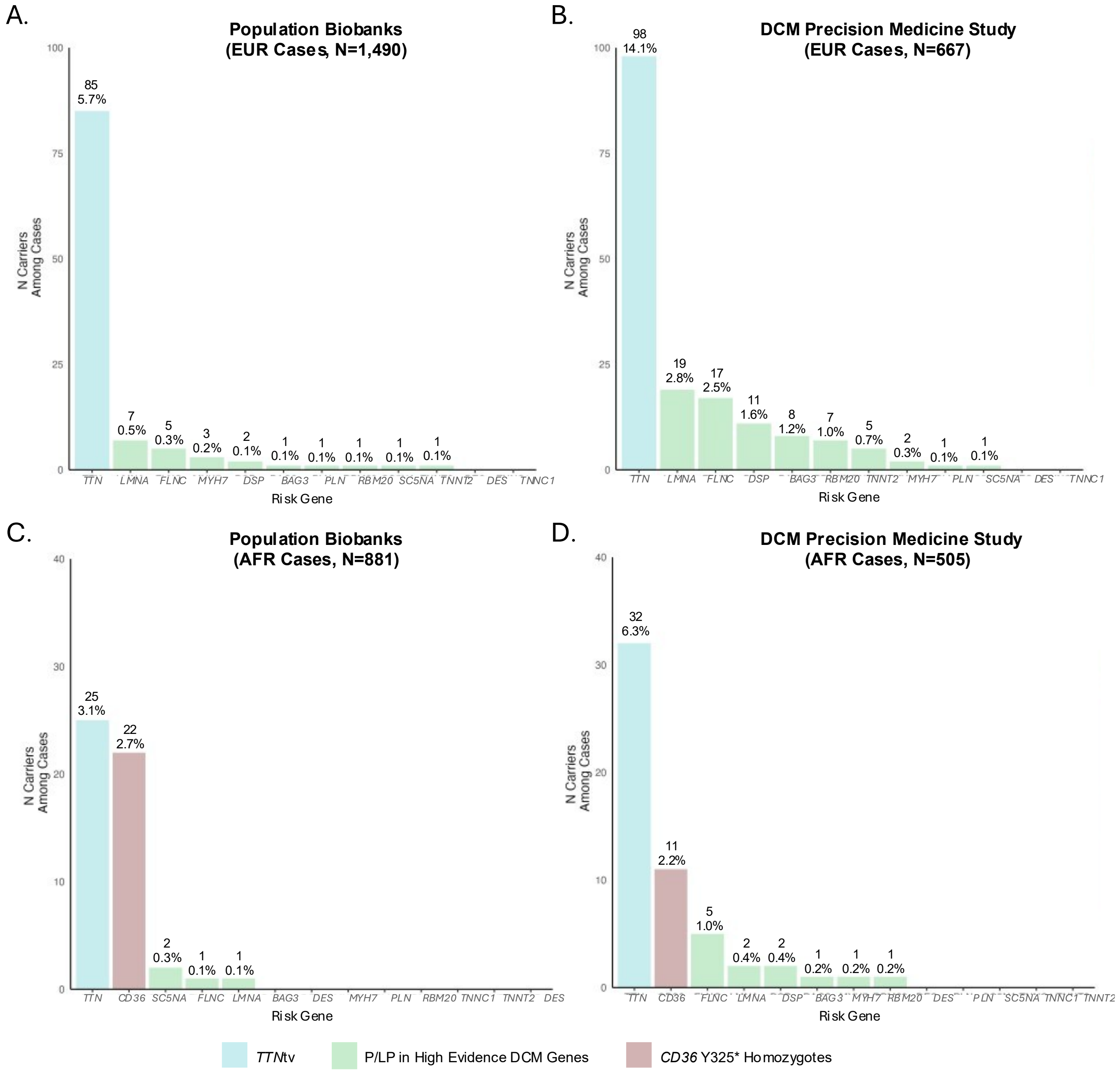

**Supplemental Figure 4: Overlap of Dilated Cardiomyopathy Risk Gene Carriers.** Figure shows the overlap of DCM risk gene carriers among 1,316 African DCM cases that originate from All of Us, Penn Medicine Biobank, Million Veteran Program, and The DCM Precision Medicine Study. The risk genes that were included in this study P/PL mutations in *TTN (TTN*tv from biobanks + clinically adjudicated *TTN* variants in DCM Precision Medicine Study), P/LP mutations in other high evidence DCM genes, *CD36* Y325* risk-allele heterozygotes, and *CD36* Y325* risk-allele homozygotes.

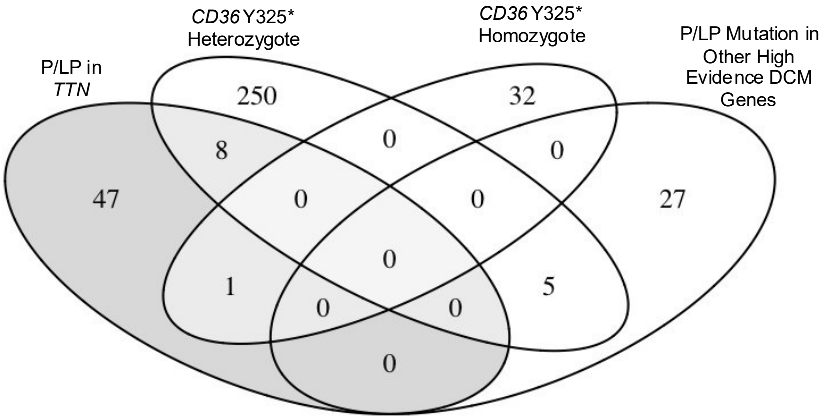

**Supplemental Figure 5: Population Attributable Fraction of Dilated Cardiomyopathy Risk Genes Across Biobank Studies by Ancestry.** Panel A illustrates the population attributable fraction for *TTNtv* and P/LP mutations in other high evidence DCM genes in European populations from All of Us, Million Veteran Program, and Penn Medicine Biobank. Panel B illustrates the population attributable fraction for *TTN*tv and *CD36* Y325* carriers in several African populations from All of Us, Million Veteran Program, and Penn Medicine Biobank.

A.

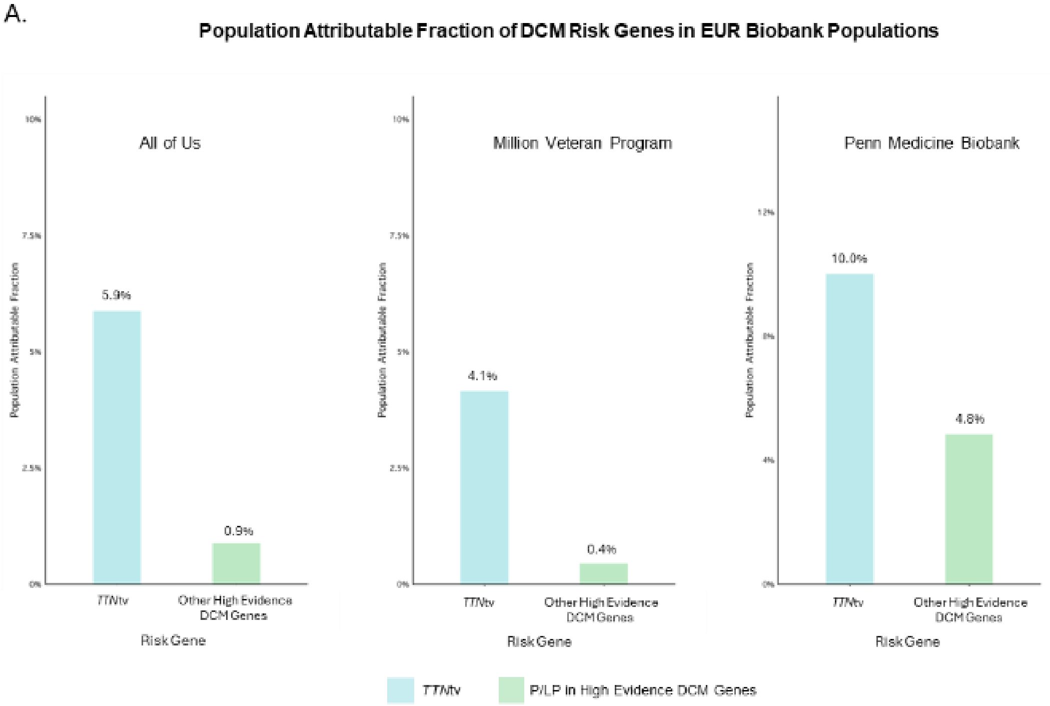


B.

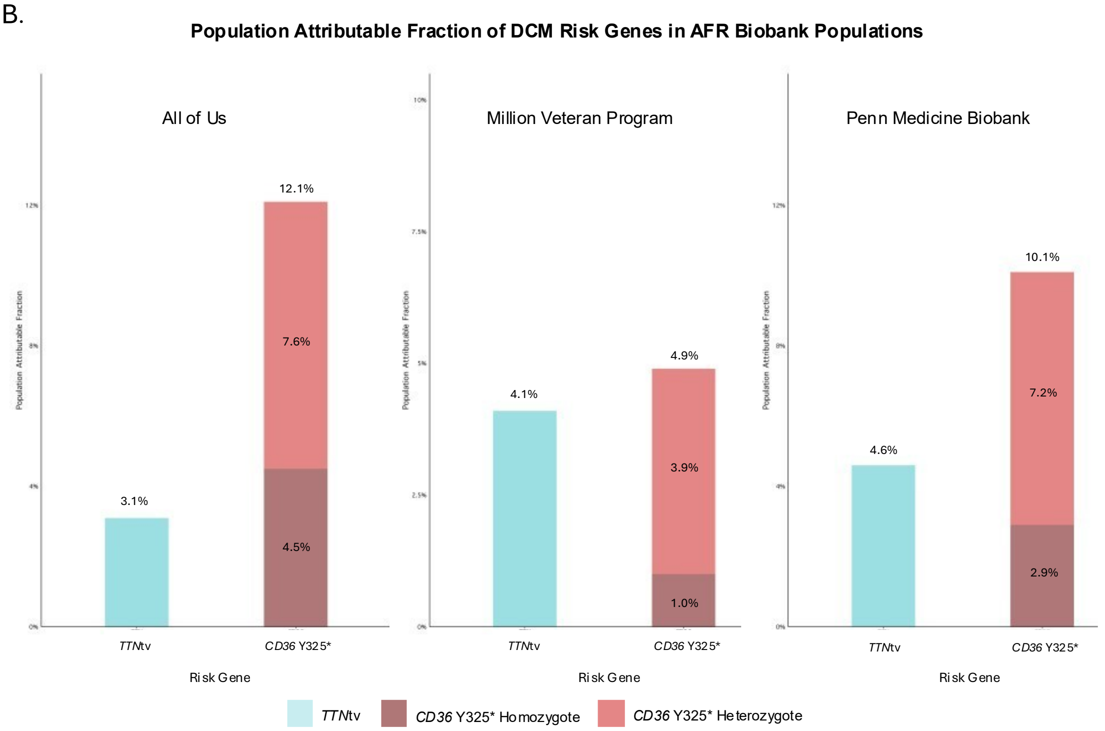

#### Supplemental Tables

Supplemental Table 1: Risk Gene Definitions.

Supplemental Table 2: Clinical Risk Factor Definitions.

Supplemental Table 3: Primary Phenotype Definitions.

Supplemental Table 4: Prevalence of DCM Risk Gene Carriers by Gene, Ancestry, and Study Recruitment Strategy.

Supplemental Table 5: Prevalence of DCM Risk Gene Carriers by Study and Ancestry.

| Risk Gene Classification | Risk Genes |
|---|---|
| **PSI90%-TTNtv** | *TTN* |
| **Other ClinGen-DCM** | *BAG3, LMNA, DSP, DES, MYH7, PLN, FLNC, RBM20, TNNC1, TNNI3, TNNT2* |

**Supplemental Table 1: Risk Gene Definitions.** This table provides a comprehensive set of risk genes and categorizes them into tiers that were used throughout the rest of this work. ClinGen-DCM genes and *TTN* represent genes that have been shown to confer outsized risk towards DCM.

| Clinical Risk Factor | ICD Codes for All of Us and Million Veteran Program | ICD Codes for Penn Medicine Biobank |
|---|---|---|
| Atrial Fibrillation | I48.X<br>[ I48.1, I48.2, I48.3, I48.4, I48.9 ] | I48.X<br>[I48.1, I48.2, I48.3, I48.4, I48.9 ] |
| Aortic Valve Disease | I06, I06.0, I06.1, I06.2, I06.8, I06.9, I08.0, I08.2, I08.3, I35, I35.0, I35.1, I35.2, I35.8, I35.9 | I06, I06.0, I06.1, I06.2, I06.8, I06.9, I08.0, I08.2, I08.3, I35, I35.0, I35.1, I35.2, I35.8, I35.9 |
| Obesity | BMI > 30 | BMI > 30 |
| Chronic Kidney Disease (CKD) | N18, I12.0, I13.1, I13.2 | N18, 11.0, 13.1, 1 |
| Chronic Obstructive Pulmonary Disease (COPD) | J41 J41.0 J42 J43 J43.0 J43.1 J43.2 J43.8 J43.9 J44 J44.0 J44.1 J44.9 491 492 496 | 491, 492, 496, J41, J41.0, J42, J43, J43.0, J43.1, J43.2 |
| Diabetes Mellitus (T2D) | E11, E11.0, E11.1, E11.2, E11.3, E11.4, E11.5, E11.5, E11.6, E11.8, E11.9 | E11 |
| Hyperlipidemia | E78.0, E78.1, E78.2, E78.4, E78.5 | E78.0, E78.1, E78.2, E78.4, E78.5 |
| Hypertension | I10 I15.9 I15.8 I15.2 I15.1 I15.0 I15 I13.2 I13.1 I13.0 I13 I12.9 I12.0 I12 I11.9 I11.0 I11 405 404 403 402 401 | I10, I10, I15.9, I15.8, I15.2, I15.1, I15.0, I15, I13.2, I13.1, I13.0, I13, I12.9, I12.0, I12, I11.9, I11.0, I11, 405, 404, 403, 402, 401 |
| Smoking Status (Ever Smoker) | Defined through questionnaire data in a study-specific fashion | Defined through questionnaire data |

**Supplemental Table 2: Clinical Risk Factor Definitions.** This table provides phenotypic constructs that were used to define several clinical risk factors in the biobank studies.

| Phenotype | ICD9 Codes | ICD10 Codes | CPT Codes |
|---|---|---|---|
| Dilated Cardiomyopathy (DCM) | NA | I42.0 | NA |
| Coronary Artery Disease (CAD) | 410, 411, 412 | I21, I22, I23, I24, I25.2 | 33510, 33511, 33512, 33513, 33533, 33534, 33535, 33536, 92920 92921, 92924, 92925, 92928, 92929, 92933, 92934, 92937, 92938, 92941, 92943, 92944, 92973, 92975 |
| Heart Failure (HF) | NA | I11.0, I13.0, I13.2, I25.4, I42, I50, O90.3 | NA |

**Supplemental Table 3: Primary Phenotype Definitions.** This table provides phenotypic constructs that were used to define the primary cardiac event phenotypes used in this study across population biobanks.

| Risk Gene | N EUR Ancestry DCM Risk Gene Carriers | | | N AFR Ancestry DCM Risk Gene Carriers | | |
|---|---|---|---|---|---|---|
| | **All DCM Cases (N = 2,157)** | **Pooled Biobank Cases (N = 1,490)** | **The DCM Precision Medicine Study Cases (N = 667)** | **All DCM Cases (N = 1,316)** | **Pooled Biobank Cases (N = 811)** | **The DCM Precision Medicine Study Cases (N = 505)** |
| *TTN* | 183 (8.5%) | 85 (5.7%) | 98 (14.7%) | 56 (4.2%) | 24 (3.0%) | 32 (6.3%) |
| *BAG3* | 9 (0.4%) | 1 (0.1%) | 8 (1.2%) | 1 (0.1%) | 0 (0%) | 1 (0.2%) |
| *DES* | 0 (0%) | 0 (0%) | 0 (0%) | 0 | 0 (0%) | 0 (0%) |
| *FLNC* | 22 (1%) | 5 (0.3%) | 17 (2.5%) | 6 (0.5%) | 1 (0.1%) | 5 (1%) |
| *LMNA* | 26 (1.2%) | 7 (0.5%) | 19 (2.8%) | 3 (0.2%) | 1 (0.1%) | 2 (0.4%) |
| *MYH7* | 5 (0.2%) | 3 (0.3%) | 2 (0.3%) | 1 (0.1%) | 0 (0%) | 1 (0.2%) |
| *PLN* | 2 (0.1%) | 1 (0.1%) | 1 (0.1%) | 0 | 0 (0%) | 0 (0%) |
| *RBM20* | 8 (0.4%) | 1 (0.1%) | 7 (1%) | 1 (0.1%) | 0 (0%) | 1 (0.2%) |
| *SC5NA* | 2 (0.1%) | 1 (0.1%) | 1 (0.1%) | 2 (0.2%) | 2 (0.3%) | 0 (0%) |
| *TNNC1* | 0 (0%) | 0 (0%) | 0 (0%) | 0 | 0 (0%) | 0 (0%) |
| *TNNT2* | 6 (0.3%) | 1 (0.1%) | 5 (0.7%) | 0 | 0 (0%) | 0 (0%) |
| *DSP* | 13 (0.6%) | 2 (0.1%) | 11 (1.6%) | 2 (0.2%) | 0 (0%) | 2 (0.2%) |
| ***CD36* Y325* Risk Allele Heterozygotes** | 0 (0%) | 0 (0%) | 0 (0%) | 267 (20.3%) | 165 (20.4%) | 102 (20.2%) |
| ***CD36* Y325* Risk Allele Homozygotes** | 0 (0%) | 0 (0%) | 0 (0%) | 33 (2.5%) | 22 (2.7%) | 11 (2.2%) |

**Supplemental Table 4: Prevalence of DCM Risk Gene Carriers by Gene, Ancestry, and Study Recruitment Strategy.** This table provides proportion of European (EUR) and African (AFR) ancestry dilated cardiomyopathy (DCM) cases that harbor pathogenic/likely pathogenic (P/LP) mutations in high evidence DCM genes, further stratified by the origin of the cases (population biobanks, recruited clinical study). The population biobanks are All of Us, Penn Medicine Biobank, and Million Veteran Program. The recruited clinical cohort is The DCM Precision Medicine Study

| | All of Us | | Million Veteran Program | | Penn Medicine Biobank | | The DCM Precision Medicine Study | |
|---|---|---|---|---|---|---|---|---|
| | **EUR** | **AFR** | **EUR** | **AFR** | **EUR** | **AFR** | **EUR** | **AFR** |
| **N Participants** | 116,624 | 48,129 | 70,836 | 23,767 | 28,980 | 10,222 | 667 | 505 |
| **N *TTN*tv** | 621 (0.5%) | 261 (0.5%) | 314 (0.4%) | 108 (0.5%) | 241 (0.8%) | 57 (0.6%) | 98 (14.7%) | 32 (6.3%) |
| **N Other ClinGen-DCM** | 101 (0.1%) | < 20 | 55 (0.1%) | < 20 | 198 (0.7%) | 35 (0.34%) | 63 (9.4%) | 10 (2.0%) |
| ***CD36* Y325* Risk Allele Heterozygotes** | **NA** | 7,834 (16.2%) | **NA** | 3,749 (15.8%) | < 20 | 1,586 (15.5%) | **NA** | 102 (20.2%) |
| ***CD36* Y325* Risk Allele Homozygotes** | **NA** | 395 (0.8%) | **NA** | 148 (0.6%) | **NA** | 87 (0.9%) | **NA** | 11 (2.2%) |

**Supplemental Table 5: Prevalence of DCM Risk Gene Carriers by Study and Ancestry.** This table provides the prevalence of dilated cardiomyopathy (DCM) risk genes among European (EUR) and African (AFR) ancestry populations within each participating study.
